# Creating and evaluating a decision support tool to reduce critically low hemoglobin and blood transfusions in-hospital

**DOI:** 10.1101/2025.09.12.25335172

**Authors:** Michael Fralick, Stephanie Lee, Meggie Debnath, Derek Beaton, Jamie Fritz, Blair Jones, Vera Dounaevskaia, Yuna Lee, Michael Colacci, Orly Bogler, Frank Rudzicz, Muhammad Mamdani

## Abstract

**Background:** Hospitalized patients have multiple risk factors for bleeding including critical illness, interventional procedures, frequent phlebotomy, and use of anticoagulant and antiplatelet medications. It is unknown if a decision support tool to identify inpatients at highest risk of bleeding can prevent critically low hemoglobin and its associated sequelae, such as requiring blood transfusion.

**Objective:** To create a decision support tool to identify hospitalized patients at high risk of bleeding sequelae (“high risk”) and to evaluate the impact of this tool on rates of critically low hemoglobin and blood transfusion.

**Methods:** We conducted a cohort study of patients hospitalized under general internal medicine at a tertiary-care teaching hospital in Toronto, Ontario. We defined “high risk” as patients with a hemoglobin < 86 g/L, or an absolute decrease in hemoglobin of ≥ 30 g/L, or a platelet count < 50 x 10^9^/L. We then created, implemented, and prospectively evaluated a decision support tool to identify these high-risk patients and alert their clinical team. Our primary outcome was the percentage of patients who received a blood transfusion.

**Results:** Our retrospective pre-deployment phase included 6,401 hospitalizations and our prospective phase included 4,274 hospitalizations. The median age of patients was 67 years (IQR 52,80), 43% were female, and the median length of stay was 5 days (IQR 2,10). Overall, 9% had a hemoglobin of 70 g/L or lower and 10% received a blood transfusion in hospital. After model implementation, the median timing of alerts was 1 day after admission (IQR 1,4). The most common trigger for an alert was a hemoglobin < 86 g/L (N=624, 76%), followed by a decrease in hemoglobin ≥ 30 g/L (N=131, 16%), and a platelet count < 50 x 10^9^/L (N=37, 5%).

Deployment of the decision support tool was associated with a reduction in the primary outcome (OR 0.78, 95% CI 0.64, 0.96), which was driven by a reduction in red blood cell transfusion (OR 0.78, 95% CI 0.64, 0.96) as opposed to platelet transfusion (OR 1.10, 95% CI 0.46, 2.91). We also observed a reduction in our secondary outcome of critically low hemoglobin (OR 0.81, 95% CI 0.65, 0.99).

**Interpretation:** Our decision support tool was associated with a modest reduction in our primary outcome of blood transfusions and secondary outcome of critically low hemoglobin. Because our study was single-centre, we believe future larger studies are needed to validate our findings.

## Introduction

One of the most common complications of being hospitalized is anemia (1,2). It affects patients on general medical wards, surgical wards, maternal wards, and in the intensive care unit. When a patient’s hemoglobin becomes critically low (i.e., approximately 70 g/L), a blood transfusion is given and often a cascade of additional investigations (e.g., additional blood work, imaging, gastroscopy) are conducted, which directly prolongs their length of stay in hospital and increases associated healthcare costs (3,4). There are multiple reasons why hospitalized patients can develop critically low hemoglobin. First, most inpatients receive daily anticoagulants for deep vein thrombosis prophylaxis (5). Second, many patients concurrently receive antiplatelet medications for comorbid conditions (e.g., coronary artery disease, stroke) or for primary prevention (6,7). Third, hospitalized patients are often acutely unwell, which adversely affects erythropoiesis and thrombopoiesis (4,8). Fourth, the median age of hospitalized adults is approximately 70 years, and anemia is common in older adults (6,9). Fifth, daily bloodwork is common while in hospital, and it contributes to iron deficiency and directly reduces patients’ iron stores, affecting their capacity to produce new red blood cells (6).

Choosing Wisely recommends minimizing daily blood work, and studies have shown that doing so can potentially improve patient outcomes and reduce costs (4,10). Despite these recommendations and potential benefits, daily blood work in hospitals remains common and causes anemia. Similarly, the use of anticoagulants for DVT prophylaxis among patients hospitalized on general medical wards is common, even though it is unnecessary for most patients and known to cause bleeding (5,11). Innovative strategies are needed to determine if preventing critically low hemoglobin might improve patient outcomes. Our objectives were to create a decision support tool to identify hospitalized patients at high risk of bleeding sequelae (“high risk”) and to evaluate the impact of this tool on rates of critically low hemoglobin and blood transfusion.

## METHODS

### Study Setting and Population

We conducted a cohort study of patients admitted to general internal medicine at St. Michael’s Hospital, a tertiary-care teaching hospital in Toronto, Ontario. Hospital services are publicly insured for residents of Ontario, and St. Michael’s Hospital serves a diverse urban population from a variety of socioeconomic backgrounds. We used data from 2019 and 2020 for exploratory analyses to develop the model and to estimate the event rate of our primary outcome. We then used retrospectively collected data (January 1, 2022 to March 31, 2023) for model development and validation, and subsequently conducted a silent deployment and clinical validation period (April 13, 2023 to August 2, 2023).

### Data Sources

Patient-level data were drawn from routinely collected data in the electronic medical record and did not require manual collection or extraction. The main categories of included data were patient demographics (e.g., age, sex), laboratory values, medications, and transfusion data. Laboratory data included hemoglobin and platelet values. Medications included anticoagulants and antiplatelets.

### Data exploration to define high-risk patients

To determine our defining criteria for patients at high risk of bleeding sequelae, we explored data from 2019 and 2020. We analyzed the distribution of hemoglobin values (including the rate of change in hemoglobin over time), platelet count, and blood transfusions. We also analyzed how the length of stay and risk of death varied across different strata of hemoglobin values to identify potential thresholds for inclusion in our model. Based on these analyses and prior literature, we selected a hemoglobin threshold of < 86 g/L. At this threshold we identified that the median length of stay for any patient with a hemoglobin < 86 g/L was 14 days, compared to 6 days for those with hemoglobin > 86 g/L. Because most patients will receive a blood transfusion if their hemoglobin falls below 70 g/L, providing an alert when the patient has a hemoglobin at least 10g/L higher gives the healthcare team time to take action to prevent further decline.

We also identified that the median rate of decline of hemoglobin over the first 72 hours was 7.0 g/L (SD 14.2). We initially chose to flag patients who had a drop in hemoglobin of 20 g/L, as this is part of the definition of major bleeding endorsed by the International Society of Thrombosis and Hemostasis (12). However, during our pilot period, we received feedback from end users that this threshold should be increased to limit false positive alarms. Thus it was raised to a ≥ 30 g/L drop in hemoglobin, which represented approximately two (i.e., 1.6) standard deviations beyond the median.

For platelet count threshold, we selected a count of < 50 x 10^9^/L as high risk, because this is a common threshold used for discontinuing full dose anticoagulants and is above the commonly used threshold for platelet transfusion of 10 x 10^9^/L (13). Our selected platelet count threshold of < 50 x 10^9^/L applies regardless of whether the patient was on an antithrombotic medication (e.g., an anticoagulant or antiplatelet). This decision was purposeful because even in the absence of an antiplatelet or anticoagulant, steps can be taken to prevent a critically low hemoglobin (e.g., stopping or reducing daily blood work, assessing for and treating iron deficiency, investigating etiology of the anemia).

In summation, we defined “high risk” as a patient with a hemoglobin < 86 g/L, or an absolute reduction of their hemoglobin of ≥ 30 g/L during the hospitalization, or a platelet count < 50 x 10^9^/L (regardless of whether the patient was on an anticoagulant or antiplatelet).

### Development of decision support tool

The decision support tool, including the aforementioned thresholds for high risk, was developed with input from the following groups: general internists, hematologists, resident physicians, and pharmacists. The initial design was adapted from a prior decision support tool we created for preventing hypoglycemia in hospital (14,15). In brief, we trained and validated a machine learning algorithm to identify patients at high-risk of bleeding using a rules-based model that adhered to the three high-risk criteria defined above.

Following the data exploration process to determine our high-risk thresholds, we used retrospectively collected data (January 1, 2022 to March 31, 2023) for model development and validation, and subsequently conducted a prospective silent deployment and clinical validation period (April 13, 2023 to August 2, 2023) to ensure the data pipelines were working correctly. Based on feedback and our prior work, our model was designed to send an email with a list of high-risk patients each day. The email was sent prior to morning rounds to ensure the relevant information was actionable during rounds. At the participating institution, each patient is assigned to a specific medical team with one most responsible physician; thus, the email was sent to the relevant team email and to the most responsible physician. Sending the email only once daily was purposeful, to limit alert fatigue. The alert was not embedded in the Electronic Medical Record (EMR) because feedback from our end-users emphasized that alerts generated by the EMR are commonly ignored, which is consistent with the available literature on alert fatigue (16).

### Deployment and iteration

We prospectively deployed the model beginning 11 September 2023. The time period from September 2023 to January 2024 is referred to as the pilot implementation. After three months of deployment, we received two primary points of feedback from end users. First, we were asked to only issue alerts for patients who had been admitted for at least 24 hours, because otherwise there is minimal lead time, and because the clinicians were often already aware of the patient’s high-risk status, as many were presenting to hospital with a gastrointestinal bleed. Second, we were asked to only send one alert per patient during their hospitalization, because multiple alerts per patient (e.g., repeating the alert the following day) was felt to be unnecessary and cumbersome. We adjusted our model in response to this feedback, making the requested changes. We launched the model for full implementation from 19 January 2024 to 20 Sep 2024.

### Study outcomes

Our primary outcome was the rate of blood transfusion defined as either receiving at least one red blood cell or platelet transfusion. Secondary outcomes included the components of the primary outcome and critically low hemoglobin (i.e., hemoglobin ≤ 70 g/L).

### Statistical Analysis

We calculated descriptive statistics to describe the study population. To assess the primary outcome, we created a logistic regression model wherein the outcome was a binary yes or no. For example, for the primary outcome of blood transfusion, “yes” included those who received a transfusion in hospital and “no” included those who did not.

## RESULTS

### Pre-deployment descriptive data

We identified 6401 hospitalizations during our pre-deployment retrospective study phase. Included patients had a median age of 67 years and 43% were female (Table 1). Across the 6401 hospitalizations, 9% of encounters had a hemoglobin ≤ 70 g/L, and 10% of encounters involved a blood transfusion, with the vast majority being red blood cell transfusions (Table 2).

**Table 1.** Baseline characteristics of included patients.

|  | **Pre-deployment**<br>**N = 6,401** | **Full implementation**<br>**N = 2,756** |
| --- | --- | --- |
| Age | 67 (52, 80) | 68 (52, 80) |
| Sex |  |  |
| Female | 2,723 (43%) | 1,254 (46%) |
| Male | 3,678 (57%) | 1,502 (54%) |
| Length of stay<br>(days) | 5 (2, 10) | 5 (2, 10) |

Legend: Numbers within brackets represent the interquartile range.

**Table 2.** Characteristics of outcomes by encounter

|  | **Pre-deployment**<br>**N = 6,401** | **Full implementation**<br>**N = 2,756** |
| --- | --- | --- |
| **Encounters with a transfusion [primary outcome]** |  |  |
| no | 5,750 (90%) | 2,510 (91%) |
| yes | 651 (10%) | 246 (9%) |
| Logistic regression | Referent | OR 0.78, 95% CI 0.64, 0.96 |
| **Encounters with a red blood cell transfusion** |  |  |
| no | 5,761 (90%) | 2,514 (91%) |
| yes | 640 (10%) | 242 (9%) |
| Logistic regression | Referent | (OR 0.78, 95% CI 0.64, 0.96) |
| **Encounters with a platelet transfusion** |  |  |
| no | 6,347 (99%) | 2,742 (99%) |
| yes | 54 (1%) | 14 (1%) |
| Logistic regression | Referent | OR 1.10, 95% CI 0.46, 2.91 |
| **Encounters with hemoglobin of 70g/L or lower** |  |  |
| no | 5,805 (91%) | 2,528 (92%) |
| yes | 596 (9%) | 228 (8%) |
| Logistic regression | Referent | OR 0.81, 95% CI 0.65, 0.99 |

### Model deployment and study outcomes

During our prospective deployment phase (which includes pilot and full implementation) from 11 September 2023 to 20 September 2024 (N=4274 hospitalizations), the median timing of alerts was 1 day after admission (IQR 1,4). The most common trigger for an alert was for a hemoglobin < 86 g/L (N=624, 76%), followed by a drop in hemoglobin of ≥ 30 g/L (N=131, 16%), and a platelet count < 50 x 10^9^/L (N=37, 5%). Antithrombotic use was common for patients with an alert (Figure 1). For example, 40% of patients with an alert had received dalteparin and 8% had received apixaban. The number of alerts per day was 2.6 (SD 1.7) during the pilot implementation period and 2.0 (SD1.4) during the full implementation period.

**Figure 1.**
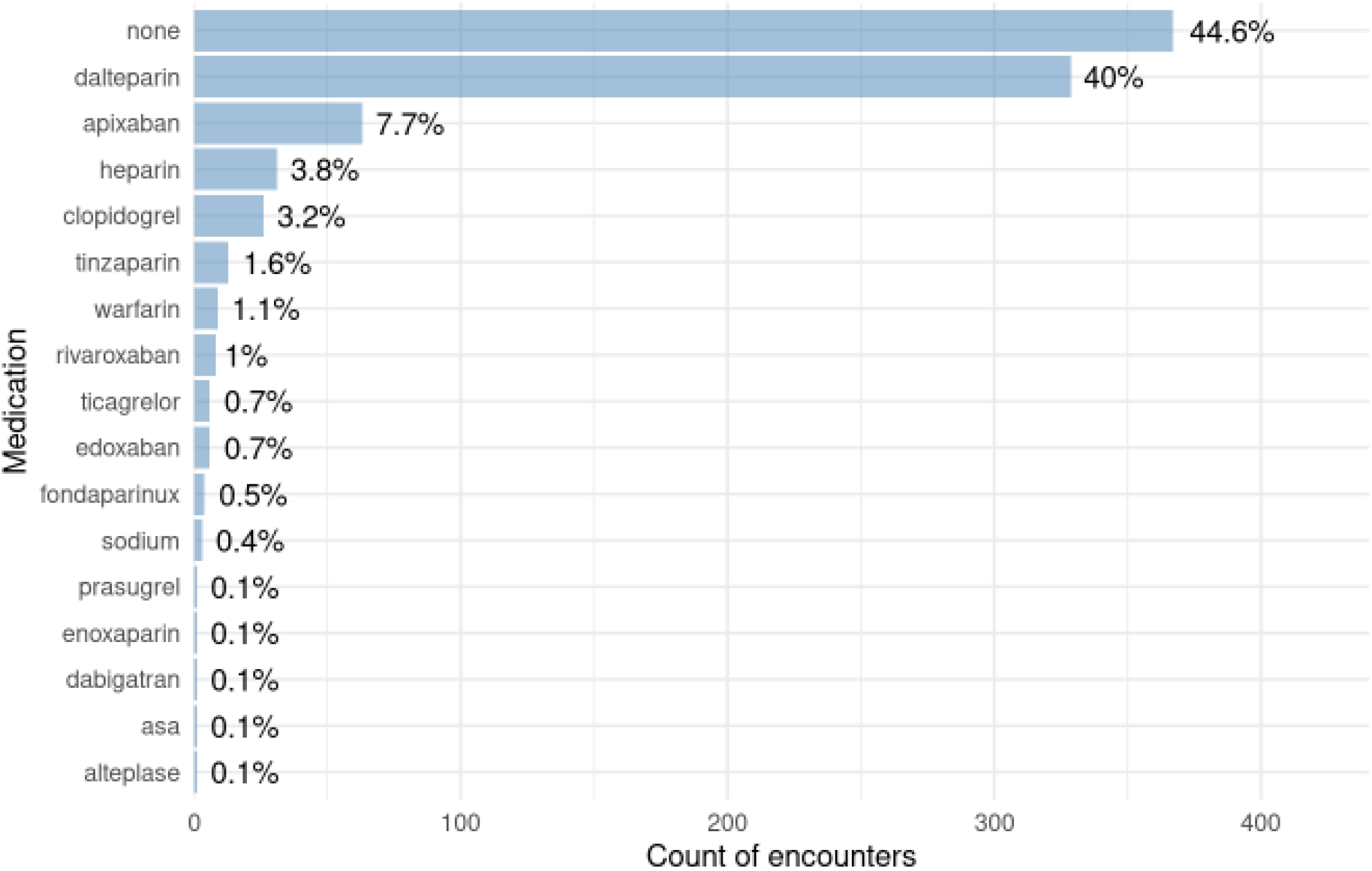
Frequency of antithrombotic medications administered among high-risk patients Legend: ASA = acetylsalicylic acid (aspirin)

For the primary outcome of inpatient transfusion, after full implementation the odds of receiving red blood cells or platelets was lower (OR 0.78, 95% CI 0.64, 0.96). The observed reduction in the primary outcome was driven by a reduction in red blood cell transfusion (OR 0.78, 95% CI 0.64, 0.96) as opposed to platelet transfusion (OR 1.10, 95% CI 0.46, 2.91). For the secondary outcome of critically low hemoglobin, we observed a modest reduction (OR 0.81, 95% CI 0.65, 0.99).

## INTERPRETATION

In this multiyear retrospective and prospective study of hospitalized patients, we were able to create, implement, and evaluate a decision support tool with the goal of reducing instances of critically low hemoglobin and blood transfusions. Our results suggest that the iterative changes we made to minimize alert fatigue resulted in a model that identified high-risk patients and led to a reduction in patients’ risk of critically low hemoglobin and blood transfusion. This observation highlights the importance of ongoing feedback from end-users to refine clinical decision support tools.

There are several strengths of our study. First, our decision support tool was tested prospectively in a general internal medicine ward, and it evaluated clinically relevant outcomes. Second, a common criticism of artificial intelligence models in healthcare is that their black box approach does not allow clinicians to know how algorithms derived their recommendations (17). Our decision support tool uses clear clinical thresholds to identify high-risk patients: a hemoglobin < 86 g/L, or a ≥ 30 g/L drop in hemoglobin, or an absolute platelet count < 50 x10^9^/L. These alerting thresholds provide clinicians with a window of opportunity to take action before the hemoglobin reaches a critical level (i.e., ≤ 70 g/L) and triggers reflexive blood transfusion and a cascade of potentially unnecessary interventions (e.g., imaging, gastroscopy). Finally, our clinical decision support tool was integrated into clinicians’ daily workflow, and it automatically extracted relevant data from the EMR without requiring any direct data input by the clinical team (18,19).

A recent meta-analysis identified four published studies that evaluated decision support tools for reducing inpatient blood transfusions (20). Following implementation, the overall odds of a patient being transfused in hospital decreased (OR 0.63, 95% CI 0.29 to 0.96) (20). The decision support tools generally provided clinicians with hemoglobin alerts at the time the clinician entered an order for a blood transfusion. While this approach is effective at decreasing inappropriate blood transfusions (e.g., ordering a blood transfusion for a patient with a hemoglobin of 100 g/L), it won’t necessarily prevent critically low hemoglobins (21–26). Our tool instead focuses on detecting high-risk patients earlier to prevent anemia progression and subsequent need for transfusion. Early detection of potential risk affords the opportunity for clinical investigation and minimization of blood loss (e.g., by stopping daily phlebotomy, or by evaluating for iron deficiency and managing accordingly).

Our study has a number of limitations. First, it is unknown how our results will apply to other populations (e.g., patients admitted under surgical services) or other hospitals. Second, we lacked data on certain bleeding risk factors (e.g., prior bleeding event, bleeding disorder, drug interactions); including these variables may have improved our model’s performance. Third, we lacked other clinically relevant outcomes such as inpatient mortality. The decision not to analyze mortality was purposeful, because our single-center study would be under-powered to analyze this outcome. Fourth, because the evaluation of our tool was observational, it is at risk of biases known to affect non-randomized studies (e.g., confounding). Finally, because our study was single-centre, future larger studies are needed to validate our findings. If similar benefits are indeed observed, our approach would provide an innovative way of reducing blood transfusions and improving inpatient care.

## Data Availability

All data generated or analyzed during this study are included in this article. Further enquiries can be directed to the corresponding author.

## Appendix

**Appendix Table 1.**
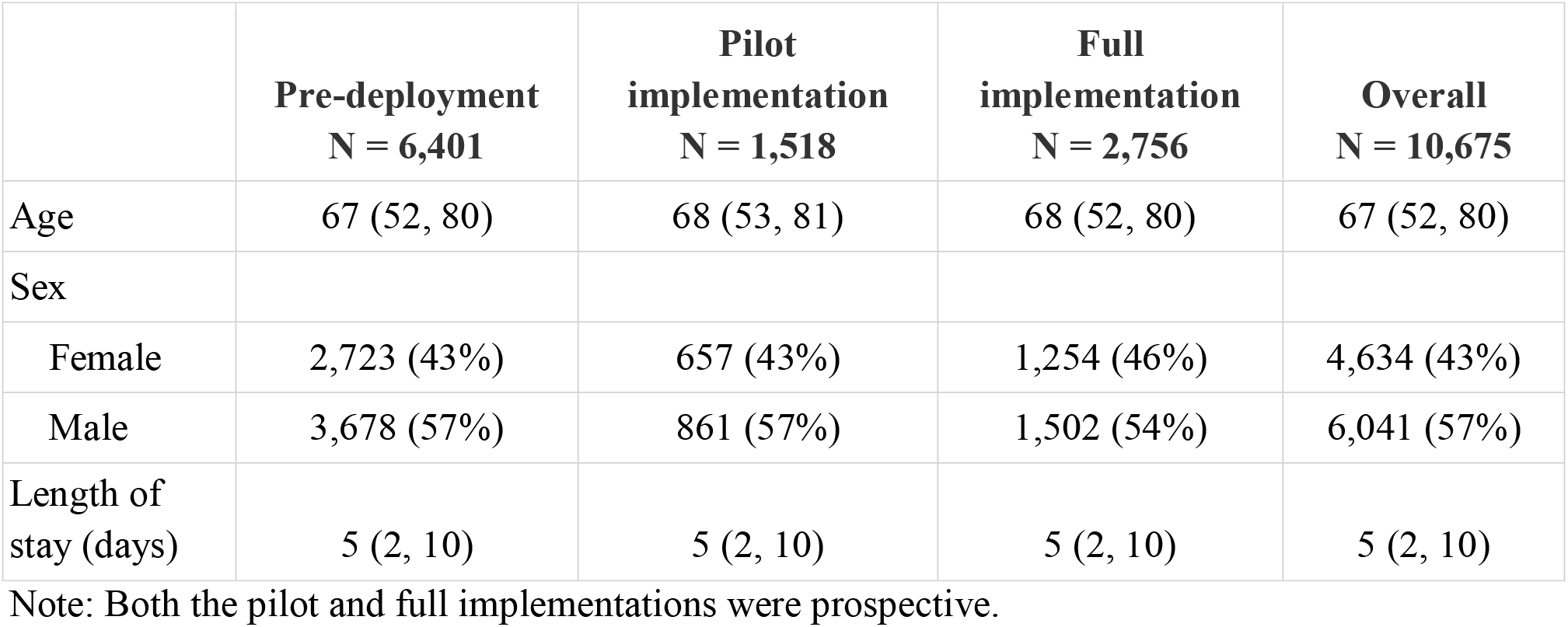
Baseline characteristics of included patients.

|  | **Pre-deployment**<br>**N = 6,401** | **Pilot**<br>**implementation**<br>**N = 1,518** | **Full**<br>**implementation**<br>**N = 2,756** | **Overall**<br>**N = 10,675** |
| --- | --- | --- | --- | --- |
| Age | 67 (52, 80) | 68 (53, 81) | 68 (52, 80) | 67 (52, 80) |
| Sex |  |  |  |  |
| Female | 2,723 (43%) | 657 (43%) | 1,254 (46%) | 4,634 (43%) |
| Male | 3,678 (57%) | 861 (57%) | 1,502 (54%) | 6,041 (57%) |
| Length of stay (days) | 5 (2, 10) | 5 (2, 10) | 5 (2, 10) | 5 (2, 10) |

Note: Both the pilot and full implementations were prospective.

**Appendix Table 2.** Characteristics of outcomes by encounter

|  | **Pre-deployment**<br>**N = 6,401** | **Pilot**<br>**implementation**<br>**N = 1,518** | **Full**<br>**implementation**<br>**N = 2,756** | **Overall**<br>**N = 10,675** |
| --- | --- | --- | --- | --- |
| Encounters with hemoglobin of 70g/L or lower |  |  |  |  |
| No | 5,805 (91%) | 1,365 (90%) | 2,528 (92%) | 9,698 (91%) |
| Yes | 596 (9%) | 153 (10%) | 228 (8%) | 977 (9%) |
| Encounters with a transfusion |  |  |  |  |
| No | 5,750 (90%) | 1,349 (89%) | 2,510 (91%) | 9,609 (90%) |
| Yes | 651 (10%) | 169 (11%) | 246 (9%) | 1,066 (10%) |
| Encounters with a red blood cell transfusion |  |  |  |  |
| No | 5,761 (90%) | 1,351 (89%) | 2,514 (91%) | 9,626 (90%) |
| Yes | 640 (10%) | 167 (11%) | 242 (9%) | 1,049 (10%) |
| Encounters with a platelet transfusion |  |  |  |  |
| No | 6,347 (99%) | 1,511 (99.5%) | 2,742 (99.5%) | 10,600 (99%) |
| Yes | 54 (1%) | 7 (0.5%) | 14 (0.5%) | 75 (1%) |

Note: Both the pilot and full implementations were prospective.

